# Saliva, a relevant alternative sample for SARS-CoV2 detection

**DOI:** 10.1101/2020.11.27.20239608

**Authors:** Monique Melo Costa, Nicolas Benoit, Jerome Dormoi, Remy Amalvict, Nicolas Gomez, Hervé Tissot-Dupont, Matthieu Million, Bruno Pradines, Samuel Granjeaud, Lionel Almeras

## Abstract

**Background:** Currently, COVID-19 diagnosis relies on quantitative reverse-transcriptase polymerase chain reaction (RT-qPCR) from nasopharyngeal swab (NPS) specimens, but NPSs present several limitations. The simplicity, low invasive and possibility of self-collection of saliva imposed this specimen as a relevant alternative for SARS-CoV-2 detection. However, the discrepancy of saliva test results compared to NPSs made of its use controversial. Here, we proposed to assess Salivettes®, as a standardized saliva collection device, and to compare SARS-CoV-2 positivity on paired NPS and saliva specimens.

**Methods:** A total of 303 individuals randomly selected among those investigated for SARS-CoV-2 were enrolled, including 30 (9.9%) patients previously positively tested using NPS (follow-up group), 90 (29.7%) mildly symptomatic and 183 (60.4%) asymptomatic.

**Results:** The RT-qPCR revealed a positive rate of 11.6% (n=35) and 17.2% (n=52) for NPSs and saliva samples, respectively. The sensitivity and specificity of saliva samples were 82.9% and 91.4%, respectively, using NPS as reference. The highest proportion of discordant results concerned the follow-up group (33.3%). Although in the symptomatic and asymptomatic groups the agreement exceeded 90.0%, 17 individuals were detected positive only in saliva samples, with consistent medical arguments.

**Conclusion:** Saliva collected with Salivette® demonstrated more sensitive for detecting symptomatic and pre-symptomatic infections.

## Introduction

In December 2019, a pneumonia outbreak caused by a novel coronavirus, Severe Acute Respiratory Syndrome Coronavirus 2 (SARS-CoV-2), emerged in Wuhan, China. Since this disease has spread quickly worldwide causing the Coronavirus Disease 2019 (COVID-19) [1]. In March 2020, this novel coronavirus has been considered as pandemic [2,3]. Breaking the chain of disease transmission was a strategy promoted by the World Health Organization (WHO) to control this health crisis. The extensive screening was recommended in order to isolate and to treat infected cases. Different biological specimens were tested for SARS-CoV-2 detection including upper and lower respiratory tracts, urinary, blood or fecal samples [4,5]. These screening tests are currently carried out by quantitative reverse transcription polymerase chain reaction (RT-qPCR) using mainly nasopharyngeal or oropharyngeal samples [6]. Although nasal/throat swabs remain the gold standard test, this collecting process causes discomfort to patients and may conduct to patient sneezing and coughing, exposing healthcare workers to viral droplets. The aerosol dispersal of SARS-CoV-2 represents a considerable risk for healthcare professionals [7]. This uncomfortable sampling method is relatively invasive and nose bleeding might also occur. Moreover, nasal swab is contraindicated for people with blood clotting diseases or deviated septum [8]. Swab sampling is technically not evident to succeed systematically, notably on very young children or self-collection [9,10]. Then, swab test sensitivity is suboptimal, and the recourse to repeat sampling is not infrequent [11].

The validation of a simple and noninvasive sampling method for SARS-CoV-2 diagnosis became on demands. Saliva was then rapidly assessed as an alternative specimen to diagnose COVID-19 [12,13]. Saliva presents numerous advantages, notably low invasive collection, easy to handle with the possibility of self-collection, permitting mass testing and exposure prevention of healthcare professionals [14]. Although reports about the screening properties accuracy of saliva for COVID-19 are more and more numerous [15–17], this biological fluid remains dispraised due to its variable diagnostic performance compared to nasopharyngeal swab (NPS) [18]. The discrepancy observed could be attributed to several factors, such as the heterogeneity of saliva specimens (eg, spitting saliva, posterior oropharyngeal, deep throat saliva, drooling collection), the saliva collection methods (eg, swabs, widemouth tube, funnel, soother), or the target population (eg, hospitalized inpatients, clinically suspected cases, symptomatic, asymptomatic, healthcare workers) [19]. To control the ongoing COVID-19 pandemic and to prevent the emergence of new foci of infection, the mass detection of symptomatic and asymptomatic individuals has become essential [20].

In this context, the present study assessed the potential of a new saliva collection system, Salivette®, for COVID-19 diagnosis. This saliva collection system, consisting to moisten a cotton roll, is hygienic, preventing saliva droplets or dripping off the collection tube. This system, dedicated mainly for cortisol measurement, could be used for drug monitoring in saliva [21,22]. The saliva collection with Salivette® was compared to conventional NPS tests using molecular assays in individuals under SARS-CoV-2 investigation. Moreover, the performances of saliva system were compared with paired NPSs among previously confirmed COVID-19 patients, symptomatic and asymptomatic individuals.

## Methods

### Ethical statement

The study protocol was reviewed and approved by the Ile de France 1 ethical committee (N°2020-A01249-30 protocol, 06/08/2020). Demographics, clinical data and samples were collected uniquely after the understanding of the study protocol and consent acknowledgement by the participants.

### Individual recruitment

During the period of 6^th^ October 2020 to 16^th^ October 2020, individuals admitted to the Institut Hospitalo-Universitaire (IHU) Méditerranée Infection (Marseille, France), for SARS-CoV-2 routine diagnosis, were invited to enroll in the research study. The inclusion criteria were all individuals on demand of SARS-CoV-2 detection using NPS, accepting in parallel, saliva collection. Individuals under 18 years old, non-French speaking, pregnant women and individuals suffering of Gougerot-Sjögren Syndrome were excluded.

### NPS management

A standard protocol was used for NPSs collection using nasal swabs with viral transport medium (Pacific Laboratory Products, Blackburn, Australia). Routine diagnosis protocol was applied for SARS-CoV-2 detection on NPS samples by RT-qPCR [23,24].

### Saliva collection

Each participant should not have eaten or drink anything in the 30 minutes prior the saliva collection. Saliva was collected using cotton roll (Neutral Salivettes®, SARSTEDT, Numbrecht, Germany) under the supervision of a laboratory technician. The cotton roll was directly introduced in the mouth without handling and then kept 2 min in the mouth’s participant who soaked the cotton by doing circular movements, prior to replace it into stopper part of the Salivette® tube. The samples were refrigerated on ice at the collection site and stored in these conditions until they arrived in the laboratory. The storing time never exceeded 6 hours.

### Saliva sample preparation

Salivettes® were centrifuged at 1500× g for 2 min at 4 °C and retrieved saliva was transferred to 1.5 mL tubes. Two hundred microliters were collected for molecular analysis and the remaining volume was preserved as rescue sample. All samples were stored at −80°C until their use. If the retrieved saliva volume was less than 150µL, 500µL of ultra-pure water were loaded at the top of the cotton roll. Salivette® was then, once again, centrifuged at 1500 × g for 2 min at 4 °C and processed as indicated above.

### RNA extraction

Viral RNA was extracted from 150 µL of the samples (NPS fluids or saliva) using NucleoMag® Pathogen Isolation kit (MACHEREY-NAGEL GmbH & Co, Düren, Germany). The nucleic acid extraction was fully automated using KingFisher™ Flex system (ThermoFisher Scientific, Villebon Courtaboeuf, France), within 28 minutes, according to the manufacturer’s instructions. The RNA was recovered in 75 μL of elution buffer and used directly as a template in RT-qPCR for SARS-CoV-2 detection.

### SARS-CoV-2 RT-Qpcr

Five microliters of eluted RNA were tested, targeting the SARS-CoV-2 envelope protein (E)-encoding gene, as previously described [24]. The Enterobacteria phage MS2 (MS2) was added to each sample as internal control [25]. All experiments were performed on a LightCycler 480II instrument (Roche Diagnostics®, Mannheim, Germany). To consider RT-qPCR, the cycle threshold (Ct) value from MS2 gene should be lower than 32 Ct. For routine diagnosis done on NPSs, samples were classified positive for SARS-CoV-2 when E primer-probe sets were detected with a Ct value < 35 [26]. The same threshold was applied to saliva samples. For the discordant results, uniquely participant classified negatives by NPSs and positives by saliva samples were informed by phone messages, and invited to perform a retest. Moreover, an examination of SARS-CoV-2 screening history was done for these participants.

### Human RNase P RT-qPCR

RT-qPCR using the Human RNase P (HRNP) primers/probe sets were performed as previously described [27], for all saliva samples in order to ensure the quality of the extraction, mainly in the samples with water addition.

### Statistical analysis

Statistical analyses were performed using the GraphPad Prism software 7.0.0 (GraphPad Software, San Diego, USA).

## Results

### Paired comparison of SARS-CoV-2 detection from NPSs and saliva samples

A total of 303 sample pairs of NPSs and saliva samples were collected. The characteristics of the participants were detailed in the Table 1. The median age was 40 years (interquartile range, 29-50; range, 18-78 years). One hundred and forty-four individuals (47.5%) were men. Concerning the symptoms, although the median onset (IQR) was 0 day (0-2.5), more than one third of the participants (n=104, 34.3%) presented symptoms at the enrolment day. The more common symptoms at presentation were fever (n=39, 12.9%), myalgia (n=31, 10.3%) and headache (n=24, 7.9%), corresponding to flu symptoms, followed by cough (n=21, 6.9%) and anosmia/ageusia (n=20, 6.6%) which were frequently reported in Covid-19 clinical diagnosis criteria [28,29].

**Table 1.** Characteristics of participants under investigation for COVID-19 diagnosis by paired NPSs and saliva samples.

|  | **Overall** | **Follow-up\*** | **Symptomatic** | **Asymptomatic** |
| --- | --- | --- | --- | --- |
| Participants, n (%) | 303 | 30 (9.9%) | 90 (29.7%) | 183 (60.4%) |
| Age (years), median (IQR) | 40 (29-50) | 45 (34.5-58.8) | 41 (28.3-50.0) | 39 (28.5-49.5) |
| Male, n (%) | 144 (47.5%) | 16 (53.3%) | 35 (38.9%) | 93 (50.8%) |
| Onset of symptoms before the test (days), median (IQR) | 0 (0-2.5) | 8 (6-11) | 3 (1-5) | / |
| Symptoms at presentation, n (%) | 104 (34.3%) | 14 (46.7%) | 90 (100%) | 0 (0.0%) |
| Cough, n (%) | 21 (6.9%) | 3 (10.0%) | 18 (20.0%) | / |
| Sore throat, n (%) | 16 (5.3%) | 1 (3.3%) | 15 (16.7%) | / |
| Runny nose, n (%) | 18 (5.9%) | 0 (0.0%) | 18 (20.0%) | / |
| Anosmia/Ageusia, n (%) | 20 (6.6%) | 6 (20.0%) | 14 (15.6%) | / |
| Diarrhea, n (%) | 6 (2.0%) | 0 (0.0%) | 6 (6.7%) | / |
| Myalgia, n (%) | 31 (10.3%) | 3 (10.0%) | 28 (31.1%) | / |
| Fever, n (%) | 39 (12.9%) | 3 (10.0%) | 36 (40.0%) | / |
| Tiredness, n (%) | 12 (4.0%) | 3 (10.0%) | 9 (10.0%) | / |
| Headache, n (%) | 24 (7.9%) | 0 (0.0%) | 24 (26.7%) | / |
| Others, n (%) | 3 (1.0%) | 1 (3.3%) | 2 (2.2%) | / |

\* Previously tested positively for SARS-CoV-2 by RT-qPCR on NPSs. Abbreviations: IQR, interquartile range; NPS, nasopharyngeal swab; SARS-CoV-2, severe acute respiratory syndrome coronavirus 2; SD, standard deviation.

Paired sample analyses revealed that the positive rate of SARS-CoV-2 screening by RT-qPCR for NPSs and saliva samples were 11.6% (n=35) and 17.2% (n=52), respectively. Among them, 29 participants were classified as positives using both specimens, and 6 and 23 were classified positively for SARS-CoV-2 only by NPSs or saliva, respectively (Table 2). The comparison of Ct values between NPSs and saliva samples were not significantly different either if all positives samples (*p*>*0*.*05*, Mann-Whitney test, Figure 1A) were considered, or if paired positive samples (*p*>*0*.*05*, Wilcoxon test, Figure 1B) were taken into account.

**Table 2.**
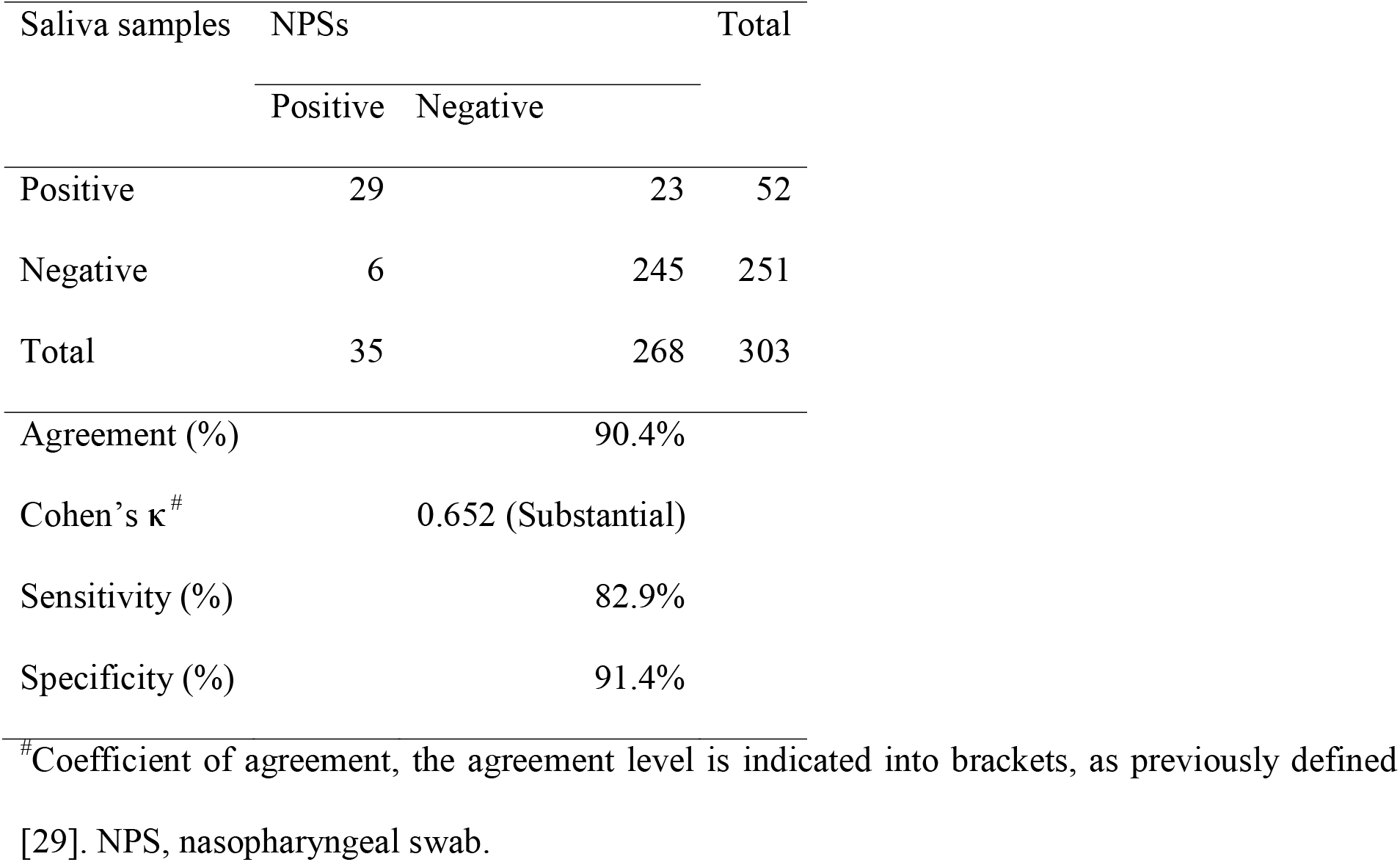
Comparison of the RT-qPCR detection of SARS-CoV-2 between NPSs and saliva samples.

**Figure 1.**
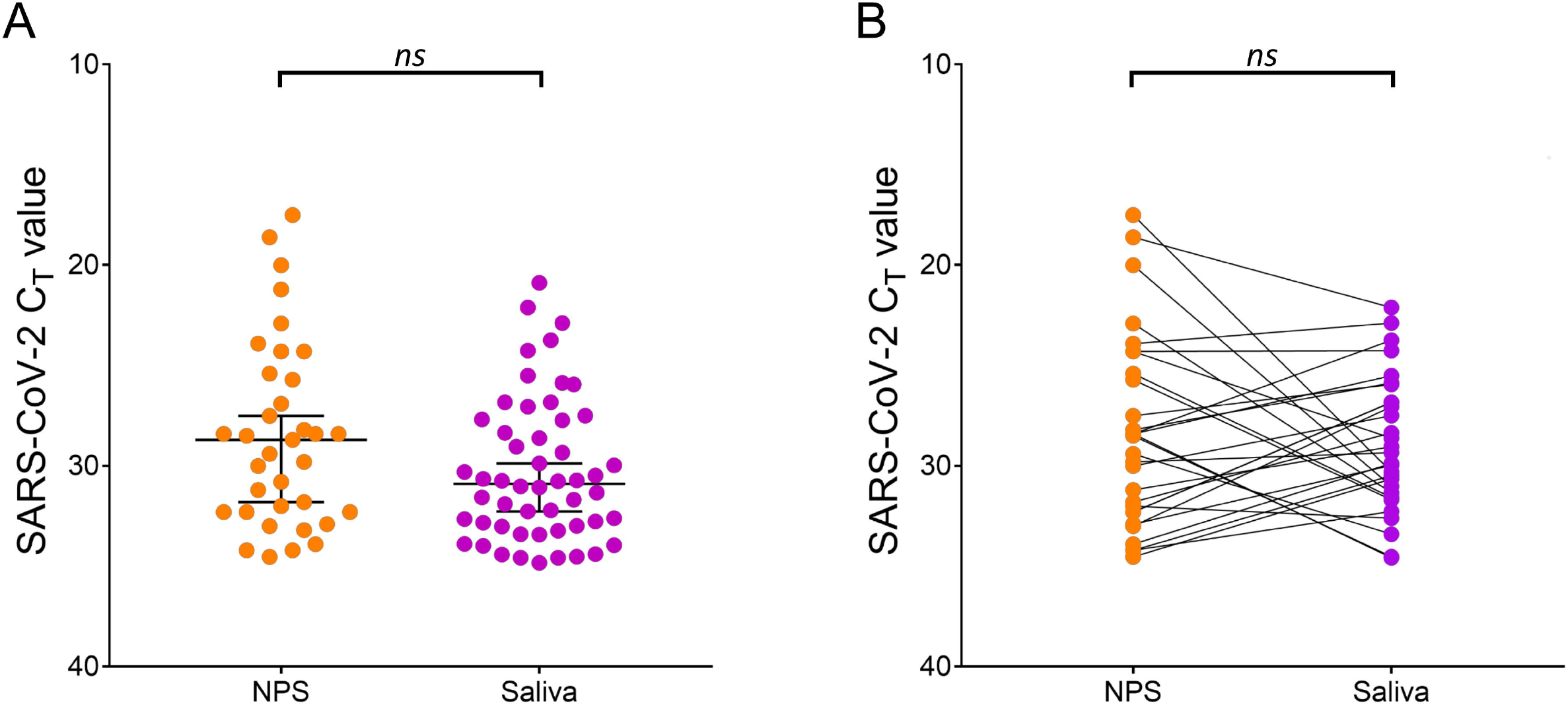
Comparison of Ct values from NPSs and Saliva samples. (A) All positive NPSs (n = 35) and saliva samples (n = 52) were compared using a Mann-Whitney test (*p = 0*.*097*). Bars represent the median and 95% CI. (B) Paired positive samples (n = 29), represented by the connecting lines, were compared by a Wilcoxon test (*p = 0*.*761*).

To determine the screening test performance of RT-qPCR on saliva samples, the results from the NPSs were used as reference. The sensitivity and specificity of saliva samples were, respectively, of 82.9% and 91.4% (Table 2). The assessment of concordance RT-qPCR results between paired specimens revealed an agreement of 90.4% with a Cohen’s κ coefficient of 0.652 corresponding to a substantial agreement of the data [30].

### Performances of screening tests according to clinical history and symptoms

Among the enrolled volunteers, 30 participants were previously positively tested for SARS-CoV-2 by RT-qPCR on NPSs and came to follow their viral load (follow-up group, n=30; 9.9%). The delay between the previous NPS SARS-CoV-2 positive tests and the present sampling ranged from 4 to 10 days. The remaining participants were separated into mildly symptomatic (n=90; 29.7%) and asymptomatic (n=183; 60.4%) groups (Table 1). No significant differences were noted between the ages (*p=0*.*189*, Kruskal-Wallis test) or the gender (*p=0*.*489*, df=2, Chi-square test) among the three groups. Conversely, the median days post-symptoms onset was significantly longer for the follow-up group compared to the symptomatic group (*p*<*0*.*001*, Mann-Whitney test). The main registered symptoms at presentation were anosmia/ageusia (n=6; 20.0%) for the follow-up group, and fever (n=36; 40.0%) and myalgia (n=28; 31.1%) for symptomatic group. The positive rate of SARS-CoV-2 screening by RT-qPCR for NPSs and saliva samples was, respectively, 20.0% (n=6) and 26.7% (n=8) for the follow-up group (*p*>*0*.*05*, Chi-square test), 24.4% (n=22) and 30.0% (n=27) for the symptomatic group (*p*>*0*.*05*, Chi-square test), and 3.8% (n=7) and 9.3% (n=17) for the asymptomatic group (*p*<*0*.*05*, Chi-square test, Table 3). In all groups, the positive rate was higher in saliva samples than in NPSs, and this difference was significantly increased for the asymptomatic group.

**Table 3.**
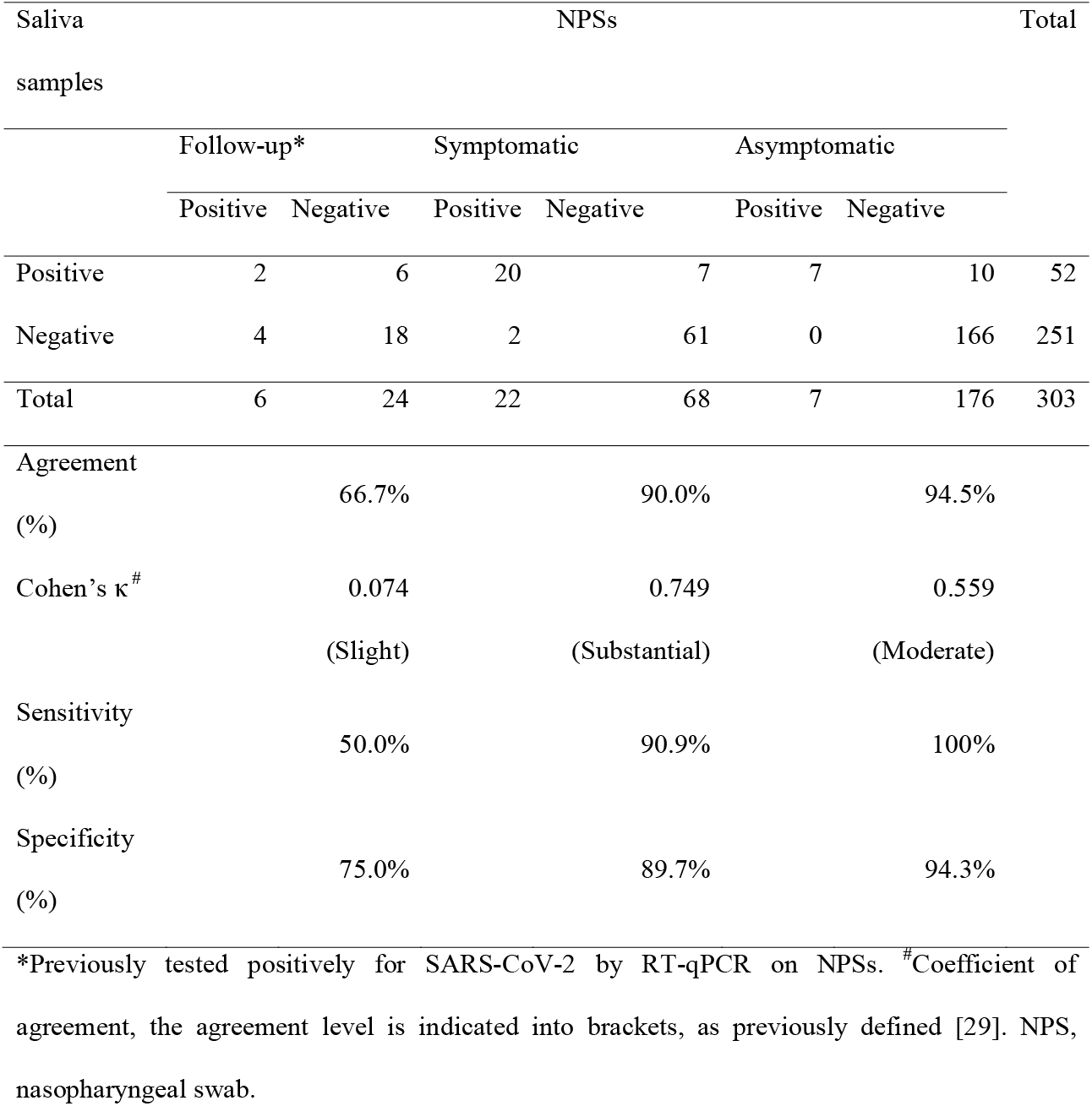
Comparison of the RT-qPCR detection of SARS-CoV-2 between NPSs and saliva samples according to participant status.

In the follow-up group (n=30), discordant results were obtained for one third (n=10) of the tested specimens leading to low sensitivity (50.0%) and specificity (75.0%) for saliva samples compared to NPSs (Table 3). Consequently, the proportion of agreement was weak (66.7%; Cohen’s κ coefficient of 0.074, slight agreement). Interestingly, among the four participants classified as positives uniquely by NPSs tests, three were detected in saliva samples with Ct values ranging from 35.1 to 36.9 (Additional File 1). Conversely, among the six participants classified as positives uniquely by saliva tests, one obtained a Ct of 36.58 using NPSs tests. Finally, only one participant was absolutely not detected by saliva specimens, whereas five were failed to be perceived by NPSs (Ct > 38).

For the symptomatic group, the sensitivity and specificity reached, respectively, 90.9% and 89.7% for saliva samples compared to NPSs (Table 3). The accuracy between the two specimens revealed an agreement of 90.0% with a Cohen’s κ coefficient of 0.749 corresponding to substantial agreement. Discordant results were obtained for nine participants. Two and seven participants were detected uniquely in NPSs and saliva samples, respectively (Additional File 1). Among the seven symptomatic participants positively tested by saliva samples, follow up information were collected for three of them. Two performed a new NPS (seven and nine days later) that confirmed their positivity (Ct values of 31.3 and 32.0). The last one indicated that his/her partner was positively detected by NPSs three days before his/her sampling. No information was collected for the remaining participants of this group.

For the asymptomatic group, a sensitivity and specificity of 100% and 94.3%, respectively, was obtained for saliva samples compared to NPSs (Table 3). The agreement proportion reached to 94.5% with a Cohen’s κ coefficient of 0.559 corresponding to moderate agreement. Ten asymptomatic participants were detected SARS-CoV-2 positive uniquely in saliva samples (Ct values ranging from 20.9 to 34.0). For three of those ten participants, flu symptoms appeared three or four days after the positive saliva test (Additional File 1). Two out of those three performed a new NPS test and were both positives (Ct values of 21.6 *vs* 32.8 three days before in saliva and 23.9 vs 20.9). Among the remaining seven of those ten participants, the partners of two participants were tested positive by NPSs two and six days before. No information was obtained from the five remaining participants positively detected by saliva samples. The analyses of clinical data and phone contacts supported that more than half of the discordant results (n=18; 62.1%) were likely true positive. The absence of complementary information from the remaining 11 participants did not allowed us to adjudicate on their infectious status.

If we consider as true positive any participants with either a NPS or a saliva positive tests, the total number of positives samples was 58, corresponding to a positive rate of 19.1% (Table 4). In these conditions, the overall proportion of agreement of SARS-CoV-2 screening for saliva (98.0%, Cohen’s κ coefficient of 0.933, almost perfect agreement), was significantly more accurate than for NPS (92.4%, Cohen’s κ coefficient of 0.711, substantial agreement; *p*<*0*.*003*, Chi-square test). The positive percent agreement (PPA, similar to sensitivity) was found significantly higher for saliva (89.7%) compared to NPS (60.3%, *p*<*0*.*003*, Chi-square test). By detecting 17 more cases than NPS, saliva obtained a clear higher performance for SARS-CoV-2 detection.

**Table 4.**
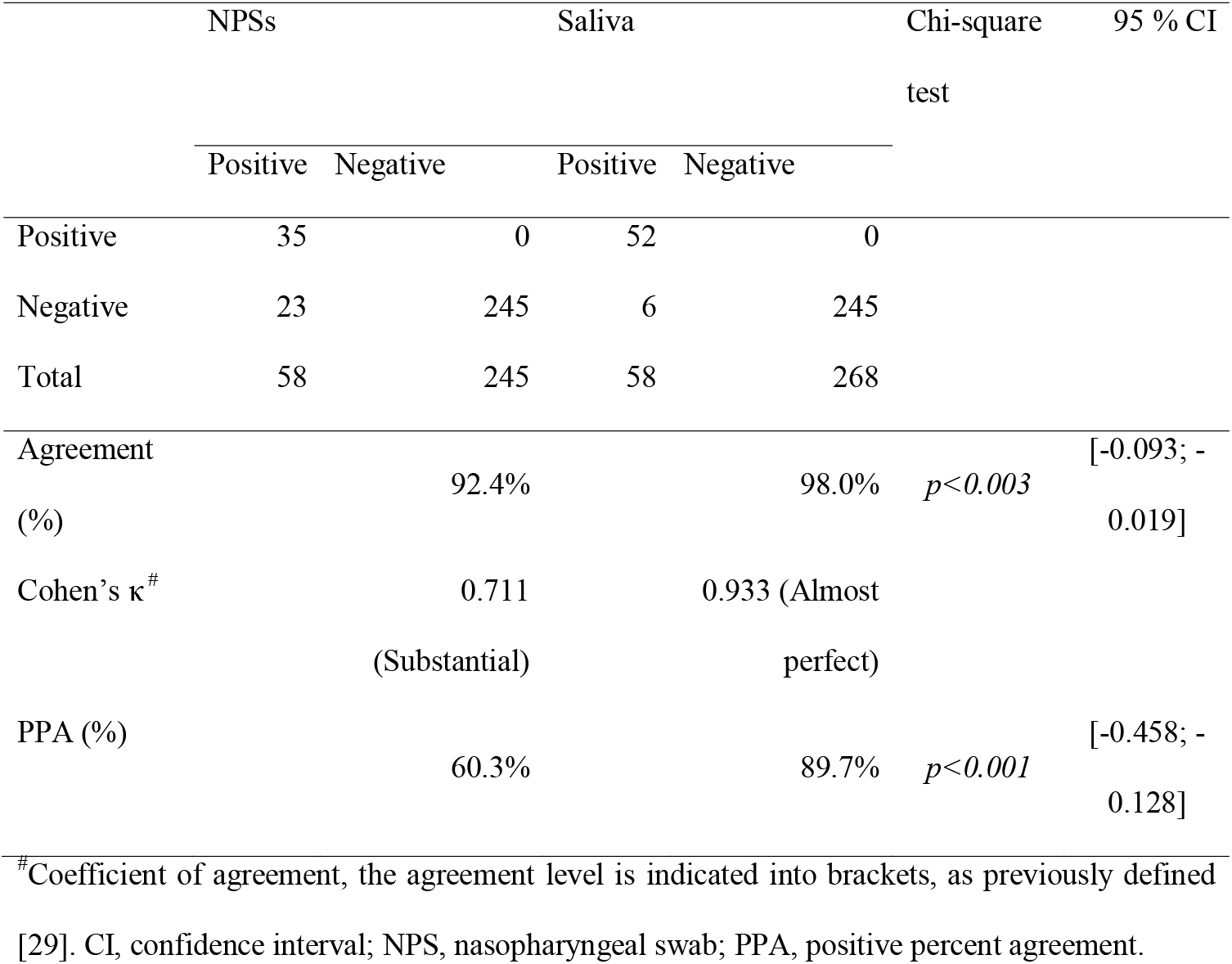
Comparison of the RT-qPCR detection of SARS-CoV-2 between NPSs and saliva samples to a reference that considers a person to be positive if one of his or her samples is positive.

|  | NPSs |  | Saliva |  | Chi-square test | 95 % CI |
| --- | --- | --- | --- | --- | --- | --- |
|  | Positive | Negative | Positive | Negative |  |  |
| Positive | 35 | 0 | 52 | 0 |  |  |
| Negative | 23 | 245 | 6 | 245 |  |  |
| Total | 58 | 245 | 58 | 268 |  |  |
| Agreement (%) | | 92.4% | | 98.0% | $p < 0.003$ | [-0.093; -0.019] |
| Cohen's $\kappa$ <sup>#</sup> | | 0.711<br>(Substantial) | | 0.933 (Almost perfect) | | |
| PPA (%) | | 60.3% | | 89.7% | $p < 0.001$ | [-0.458; -0.128] |

<sup>#</sup>Coefficient of agreement, the agreement level is indicated into brackets, as previously defined [29]. CI, confidence interval; NPS, nasopharyngeal swab; PPA, positive percent agreement.

### RNA detection in diluted saliva samples

To control whether the addition of ultra-pure water to saliva samples could be detrimental for RNA detection, a comparison of Ct values from HRNP between the 34 saliva samples with addition of ultra-pure water, and 269 remaining saliva samples was performed. No HRNP amplification was obtained for one saliva sample with water addition, whereas, the failing of PCR products in four saliva samples, which did not required water addition, was observed. HRNP Ct values from saliva samples with water addition were significantly lower than those without water addition (*p*<*0*.*003*, Mann-Whitney test, Additional File 2A). Nevertheless, among the 34 saliva samples filled up with ultra-pure water, SARS-CoV-2 was detected in five samples (Ct values ranging from 29.9 to 34.0). HRNP products were obtained for these five samples (Ct values ranging from 26.7 to 33.2). Among them, one participant, positively detected for SARS-CoV-2 using saliva (Ct value: 29.9), was also corroborated by the NPS specimen (Ct value: 33.9). These results indicated that, despite water addition decreased detection of human RNA cellular control, SARS-CoV-2 detection was not compromised by saliva dilution. The non-access of NPS specimens did not allowed to assess HRNP detection, and then to conclude whether miss-detection in NPSs was attributed to improper sampling (no PCR product) or to insufficient local viral titers (lower than detection limit).

Interestingly, the HRNP was detected in the 52 SARS-CoV-2 saliva samples and no significant difference of HRNP Ct values was observed between positives and negatives SARS-CoV-2 saliva samples (*p*>*0*.*05*, Mann-Whitney test, Additional File 2B). Moreover, HRNP was detected in the 6 saliva samples (Ct values ranging from 29.9 to 33.9), for which respective NPS specimens were found SARS-CoV-2 positives. The miss-detection of the coronavirus in these saliva samples was not due to RNA extraction impairment.

## Discussion

In the present study, Salivette® devices were chosen to standardize saliva collection. For an accurate comparison of saliva and NPS specimens, the same extraction method and RT-qPCR systems, with identical experiment control and validation criteria, were used. Overall, the present study confirmed the relevance of the saliva sampling for SARS-CoV-2 detection in the 303 individuals under investigations, with a concordance to NPSs exceeding 90%, that was among the more efficient [19]. Nevertheless, among the 58 participants classified SARS-CoV-2 positive either by NPS and/or saliva tests, discordant paired detection represented half of them (n=29). Most of the discordant pairs were detected positive only in saliva samples (79.3%, 23/29). Investigations based on clinical history analyses and patient phone contacts revealed a confirmation or supportive data of SARS-CoV-2 infection for 60.9% of them (14/23). The lack of data hampered to conclude on the SARS-CoV-2 *status* of the remaining participants. If any participant with either a NPS or a saliva positive tests is considered a true positive, as it is frequently done [31,32], the PPA was incontestably significantly higher for saliva than in NPS, which was consistent with previous works reporting an increased sensitivity of saliva specimens [33–36]. The absence of significant differences in SARS-CoV-2 Ct values, when all positives or paired positive specimens were compared, confirms that the increased sensitivity could not be attributed to an improved detection of the virus in saliva. The inconsistency of NPS sampling that induces false negatives seems to be a recurrent phenomenon especially in patients with a low viral load [37], frequently reported during serial testing [11,38,39] or in early stage of infection [40].

To limit false negative detection, RNA template or synthetic RNA were proposed as external controls for the proper extraction and amplification [41]. However, these external RNAs did not insure a proper sample collection nor the integrity of the RNA sample [24,27,42], and the use of a human RNA cellular control was proposed. The US CDC proposed to use HRNP as a control of proper sampling, sample preservation and extraction [43]. In our study, we did not access to the NPS samples of the routine diagnosis, then it was no possible to control whether the discordance between NPS and saliva specimens was attributed to a RNA integrity failure or a miss-detection of the virus. Conversely, the amplification of the HRNP in the saliva samples from participants classified positive by NPS tests, underlined that the miss-detection seems more attributed to an absence or a sub-detection of the virus rather a problem of saliva collection. The systematic used of human RNA cellular control is mandatory whatever the sampling sources to improve results interpretation.

Although, the risk to fail saliva sampling seems less frequent than for NPSs, in a recent study assessing the efficiency of Salivette® for screening SARS-CoV-2 hospitalized cases, 12.2% (6/49) of patients were excluded for lack of enough saliva volume [32]. The sialadenitis, an acute inflammation of salivary glands reported in COVID-19 patients, could conduct to a decrease of salivary flow compromising saliva collection [44]. We observed an insufficient volume of saliva in 11.2% of the samples (n=34). The large majority of these individuals (29/34; 85.3%) were tested SARS-CoV-2 negative in saliva, suggesting that, here, lower salivation could not be attributed to viral infection. Concerning the human cellular RNA control, adding water did not impair the detection of the HRNP. Although significant differences of HRNP Ct values were noticed between diluted and not diluted saliva samples, the SARS-CoV-2 detection in five saliva specimens underlined that viral detection seems not altered by diluting saliva. The water addition at the top of cotton roll allowed to recover most of the saliva samples (n=33/34, 97.1%), for which HRNP was detected. Finally, water addition did not compromised RNA detection and the RNA integrity control allows to reduce false negative detection.

The saliva tests were also compared to NPSs according to clinical history and symptomatology of individuals. A weak agreement was observed in the follow-up group. As patients from this group were convalescent, their viral charge is decreasing [12], and then miss-paired viral detection could occurred. Among them ten discordant detection, four could be considered as non-contagious based on Ct values from paired samples (35<Ct<38) [26]. The decrease in viral load after the first week subsequently the onset of symptoms is consistent with previous findings in NPSs [45,46], as well as in saliva samples [47,48].

To contain the COVID-19 spreading, an accurate identification of infectious patients regarding SARS-CoV-2 among symptomatic and asymptomatic individuals is essential. Here, although proportion of agreement between NPS and saliva specimens exceeded 90%, numerous individuals (n=17) were detected only in saliva samples. Saliva collected with Salivette® appeared more sensitive for detecting symptomatic and pre-symptomatic infections. Complementary experiments in others routine diagnosis sites are required to validate these results. Previously, the used of Salivette® devices on COVID-19 hospitalized patients reported a sensibility and specificity of 100% and 97.2%, respectively, when NPSs were used as references [32]. Salivette® presents several advantages, such as a better acceptation than NPSs for sample collection, notably for children. Moreover, the inoffensiveness of sampling allows to perform daily collection from hospitalized patients to follow viral titer.

## Conclusion

Europa is heading into the winter months and the risk of influenza symptoms is expected to increase that which should make difficult to distinct COVID-19 from non-COVID-19 individuals based on clinical symptoms. The setup of a screening system allowing rapid self-collection and accurate SARS-CoV-2 detection should be helpful in isolating infectious individuals. The present work confirmed saliva test as a reliable alternative to NPSs for SARS-CoV-2 detection. Saliva specimens showed significantly higher performance for detection of the virus in symptomatic and asymptomatic individuals. The use of a standardized saliva collection device and routine sample integrity testing with a human cellular RNA should reduce inter-sample variation and this combination should validate the use of saliva as a diagnostic tool for SARS-CoV-2. A short-time validation of these guidelines by another SARS-CoV-2 screening center became compulsory.

## Data Availability

All the data were included in the manuscript

## List of abbreviations

NPS: nasopharyngeal swab
PCR: Polymerase Chain Reaction
Ct: Cycle threshold
RT-qPCR: Reverse transcription quantitative real-time PCR
SARS-CoV-2: Severe Acute Respiratory Syndrome Coronavirus 2
COVID-19: Coronavirus Disease 2019
PPA: positive percent agreement

## Authors’ contributions

Conceived and designed the experiments: LA. Performed the experiments: MMC, NB, LA. Analyzed the data: LA, SG, MMC. Contributed reagents/materials/analysis tools: MMC, SG, JD, RA, NG, HTD, MM. Sample collections: LA, JD, RA, NG. Drafted the paper: LA. Revised critically the paper: all the authors.

## Acknowledgments

We would like to acknowledge IHU Mediterranean Infection nurse staff for their reception in the routine test rooms and all the participants who accepted to donate saliva. We also acknowledge Catherine Verret and Carine Malle (DFRI, SSA, Paris) for their help in the redaction of the ethical statement folder.

## Supporting information files

**Additional File 1. SARS-CoV-2 Ct values from discordant results of paired NPS and saliva samples according to clinical history or symptoms**. Paired samples with Ct>38 were not presented. Paired samples detected at a non-infectious level (35<Ct<38) are indicated by squares. Triangles represent participants with flu symptoms apparition with (arrow up) or without (arrow down) confirmed SARS-CoV-2 positive test. The participants for who his/her partner was declared SARS-CoV-2 positive few days before are represented by diamond.

**Additional File 2. Consequences of water addition to saliva samples on RNA detection**. (A) Comparison of human RNase P Ct values between saliva samples with (n=34) and without (n=269) water addition (*p*<*0*.*003*, Mann-Whitney test). (B) Comparison of human RNase P Ct values between positive (n=52) and negative (n=251) SARS-Cov-2 saliva samples (*p*>*0*.*05*, Mann-Whitney test). Bars represent the median and 95% CI.

## References

1. Li Q, Guan X, Wu P, et al. Early Transmission Dynamics in Wuhan, China, of Novel Coronavirus-Infected Pneumonia. N Engl J Med. 2020; 382(13):1199–1207.

2. Astuti I, Ysrafil null. Severe Acute Respiratory Syndrome Coronavirus 2 (SARS-CoV-2): An overview of viral structure and host response. Diabetes Metab Syndr. 2020; 14(4):407–412.

3. Thompson R. Pandemic potential of 2019-nCoV. Lancet Infect Dis. 2020; 20(3):280.

4. Wang W, Xu Y, Gao R, et al. Detection of SARS-CoV-2 in Different Types of Clinical Specimens. JAMA. 2020; 323(18):1843–1844.

5. Lai CKC, Lam W. Laboratory testing for the diagnosis of COVID-19. Biochem Biophys Res Commun. 2020;.

6. Hussein HA, Hassan RYA, Chino M, Febbraio F. Point-of-Care Diagnostics of COVID-19: From Current Work to Future Perspectives. Sensors. 2020; 20(15).

7. Scheuch G. Breathing Is Enough: For the Spread of Influenza Virus and SARS-CoV-2 by Breathing Only. J Aerosol Med Pulm Drug Deliv. 2020; 33(4):230–234.

8. Sri Santosh T, Parmar R, Anand H, Srikanth K, Saritha M. A Review of Salivary Diagnostics and Its Potential Implication in Detection of Covid-19. Cureus. 2020; 12(4):e7708.

9. Liao W-T, Hsu M-Y, Shen C-F, Hung K-F, Cheng C-M. Home Sample Self-Collection for COVID-19 Patients. Adv Biosyst. 2020;:e2000150.

10. Yee R, Truong T, Pannaraj PS, et al. Saliva is a promising alternative specimen for the detection of SARS-CoV-2 in children and adults. MedRxiv Prepr Serv Health Sci. 2020;.

11. Zou L, Ruan F, Huang M, et al. SARS-CoV-2 Viral Load in Upper Respiratory Specimens of Infected Patients. N Engl J Med. 2020; 382(12):1177–1179.

12. To KK-W, Tsang OT-Y, Chik-Yan Yip C, et al. Consistent detection of 2019 novel coronavirus in saliva. Clin Infect Dis Off Publ Infect Dis Soc Am. 2020;.

13. Thompson RN, Cunniffe NJ. The probability of detection of SARS-CoV-2 in saliva. Stat Methods Med Res. 2020; 29(4):1049–1050.

14. Sapkota D, Søland TM, Galtung HK, et al. COVID-19 salivary signature: diagnostic and research opportunities. J Clin Pathol. 2020;.

15. Yokota I, Shane PY, Okada K, et al. Mass screening of asymptomatic persons for SARS-CoV-2 using saliva. Clin Infect Dis Off Publ Infect Dis Soc Am. 2020;.

16. Fakheran O, Dehghannejad M, Khademi A. Saliva as a diagnostic specimen for detection of SARS-CoV-2 in suspected patients: a scoping review. Infect Dis Poverty. 2020; 9(1):100.

17. Senok A, Alsuwaidi H, Atrah Y, et al. Saliva as an Alternative Specimen for Molecular COVID-19 Testing in Community Settings and Population-Based Screening. Infect Drug Resist. 2020; 13:3393– 3399.

18. Czumbel LM, Kiss S, Farkas N, et al. Saliva as a Candidate for COVID-19 Diagnostic Testing: A Meta-Analysis. Front Med. 2020; 7:465.

19. Comber L, Walsh KA, Jordan K, et al. Alternative clinical specimens for the detection of SARS-CoV-2: A rapid review. Rev Med Virol. 2020;.

20. Li R, Pei S, Chen B, et al. Substantial undocumented infection facilitates the rapid dissemination of novel coronavirus (SARS-CoV-2). Science. 2020; 368(6490):489–493.

21. Losiak W, Losiak-Pilch J. Cortisol Awakening Response, Self-Reported Affect and Exam Performance in Female Students. Appl Psychophysiol Biofeedback. 2020; 45(1):11–16.

22. Soderstrom JH, Fatovich DM, Mandelt C, et al. Correlation of paired toxic plasma and saliva paracetamol concentrations following deliberate self-poisoning with paracetamol. Br J Clin Pharmacol. 2012; 74(1):154–160.

23. Amrane S, Tissot-Dupont H, Doudier B, et al. Rapid viral diagnosis and ambulatory management of suspected COVID-19 cases presenting at the infectious diseases referral hospital in Marseille, France, - January 31st to March 1st, 2020: A respiratory virus snapshot. Travel Med Infect Dis. 2020; 36:101632.

24. Corman VM, Landt O, Kaiser M, et al. Detection of 2019 novel coronavirus (2019-nCoV) by real-time RT-PCR. Euro Surveill Bull Eur Sur Mal Transm Eur Commun Dis Bull. 2020; 25(3).

25. Ninove L, Nougairede A, Gazin C, et al. RNA and DNA bacteriophages as molecular diagnosis controls in clinical virology: a comprehensive study of more than 45,000 routine PCR tests. PloS One. 2011; 6(2):e16142.

26. La Scola B, Le Bideau M, Andreani J, et al. Viral RNA load as determined by cell culture as a management tool for discharge of SARS-CoV-2 patients from infectious disease wards. Eur J Clin Microbiol Infect Dis Off Publ Eur Soc Clin Microbiol. 2020; 39(6):1059–1061.

27. Vogels CBF, Brito AF, Wyllie AL, et al. Analytical sensitivity and efficiency comparisons of SARS-CoV-2 RT-qPCR primer-probe sets. Nat Microbiol. 2020; 5(10):1299–1305.

28. Al-Ani RM, Acharya D. Prevalence of Anosmia and Ageusia in Patients with COVID-19 at a Primary Health Center, Doha, Qatar. Indian J Otolaryngol Head Neck Surg Off Publ Assoc Otolaryngol India. 2020;:1–7.

29. Qiu C, Cui C, Hautefort C, et al. Olfactory and Gustatory Dysfunction as an Early Identifier of COVID-19 in Adults and Children: An International Multicenter Study. Otolaryngol--Head Neck Surg Off J Am Acad Otolaryngol-Head Neck Surg. 2020; 163(4):714–721.

30. Landis JR, Koch GG. The measurement of observer agreement for categorical data. Biometrics. 1977; 33(1):159–174.

31. Migueres M, Mengelle C, Dimeglio C, et al. Saliva sampling for diagnosing SARS-CoV-2 infections in symptomatic patients and asymptomatic carriers. J Clin Virol Off Publ Pan Am Soc Clin Virol. 2020; 130:104580.

32. Aita A, Basso D, Cattelan AM, et al. SARS-CoV-2 identification and IgA antibodies in saliva: One sample two tests approach for diagnosis. Clin Chim Acta Int J Clin Chem. 2020; 510:717–722.

33. Wyllie AL, Fournier J, Casanovas-Massana A, et al. Saliva or Nasopharyngeal Swab Specimens for Detection of SARS-CoV-2. N Engl J Med. 2020; 383(13):1283–1286.

34. Jamal AJ, Mozafarihashjin M, Coomes E, et al. Sensitivity of nasopharyngeal swabs and saliva for the detection of severe acute respiratory syndrome coronavirus 2 (SARS-CoV-2). Clin Infect Dis Off Publ Infect Dis Soc Am. 2020;.

35. Ali F, Sweeney DA. No One Likes a Stick up Their Nose: Making the Case for Saliva-Based Testing for COVID-19. Clin Infect Dis Off Publ Infect Dis Soc Am. 2020;.

36. Rao M, Rashid FA, Sabri FSAH, et al. Comparing nasopharyngeal swab and early morning saliva for the identification of SARS-CoV-2. Clin Infect Dis Off Publ Infect Dis Soc Am. 2020;.

37. Petrillo S, Carrà G, Bottino P, et al. A Novel Multiplex qRT-PCR Assay to Detect SARS-CoV-2 Infection: High Sensitivity and Increased Testing Capacity. Microorganisms. 2020;8(7).

38. Wölfel R, Corman VM, Guggemos W, et al. Virological assessment of hospitalized patients with COVID-2019. Nature. 2020; 581(7809):465–469.

39. Zhao J, Yuan Q, Wang H, et al. Antibody responses to SARS-CoV-2 in patients of novel coronavirus disease 2019. Clin Infect Dis Off Publ Infect Dis Soc Am. 2020;.

40. Xie X, Zhong Z, Zhao W, Zheng C, Wang F, Liu J. Chest CT for Typical Coronavirus Disease 2019 (COVID-19) Pneumonia: Relationship to Negative RT-PCR Testing. Radiology. 2020; 296(2):E41–E45.

41. Wang D, Hu B, Hu C, et al. Clinical Characteristics of 138 Hospitalized Patients With 2019 Novel Coronavirus-Infected Pneumonia in Wuhan, China. JAMA. 2020; 323(11):1061–1069.

42. Chu DKW, Pan Y, Cheng SMS, et al. Molecular Diagnosis of a Novel Coronavirus (2019-nCoV) Causing an Outbreak of Pneumonia. Clin Chem. 2020; 66(4):549–555.

43. CDC. Coronavirus Disease 2019 (COVID-19). Centers for Disease Control and Prevention [Internet]. Available from: https://www.cdc.gov/coronavirus/2019-ncov/lab/rt-pcr-panel-primer-probes.html (2020).

44. Wang C, Wu H, Ding X, et al. Does infection of 2019 novel coronavirus cause acute and/or chronic sialadenitis? Med Hypotheses. 2020; 140:109789.

45. Wang Y, Zhang L, Sang L, et al. Kinetics of viral load and antibody response in relation to COVID-19 severity. J Clin Invest. 2020; 130(10):5235–5244.

46. Liu Y, Yan L-M, Wan L, et al. Viral dynamics in mild and severe cases of COVID-19. Lancet Infect Dis. 2020; 20(6):656–657.

47. Zhu J, Guo J, Xu Y, Chen X. Viral dynamics of SARS-CoV-2 in saliva from infected patients. J Infect. 2020; 81(3):e48–e50.

48. To KK-W, Tsang OT-Y, Leung W-S, et al. Temporal profiles of viral load in posterior oropharyngeal saliva samples and serum antibody responses during infection by SARS-CoV-2: an observational cohort study. Lancet Infect Dis. 2020; 20(5):565–574.

