## Supplementary figures and images for "Saliva, a relevant alternative sample for SARS-CoV2 detection"

### Additional_File-1

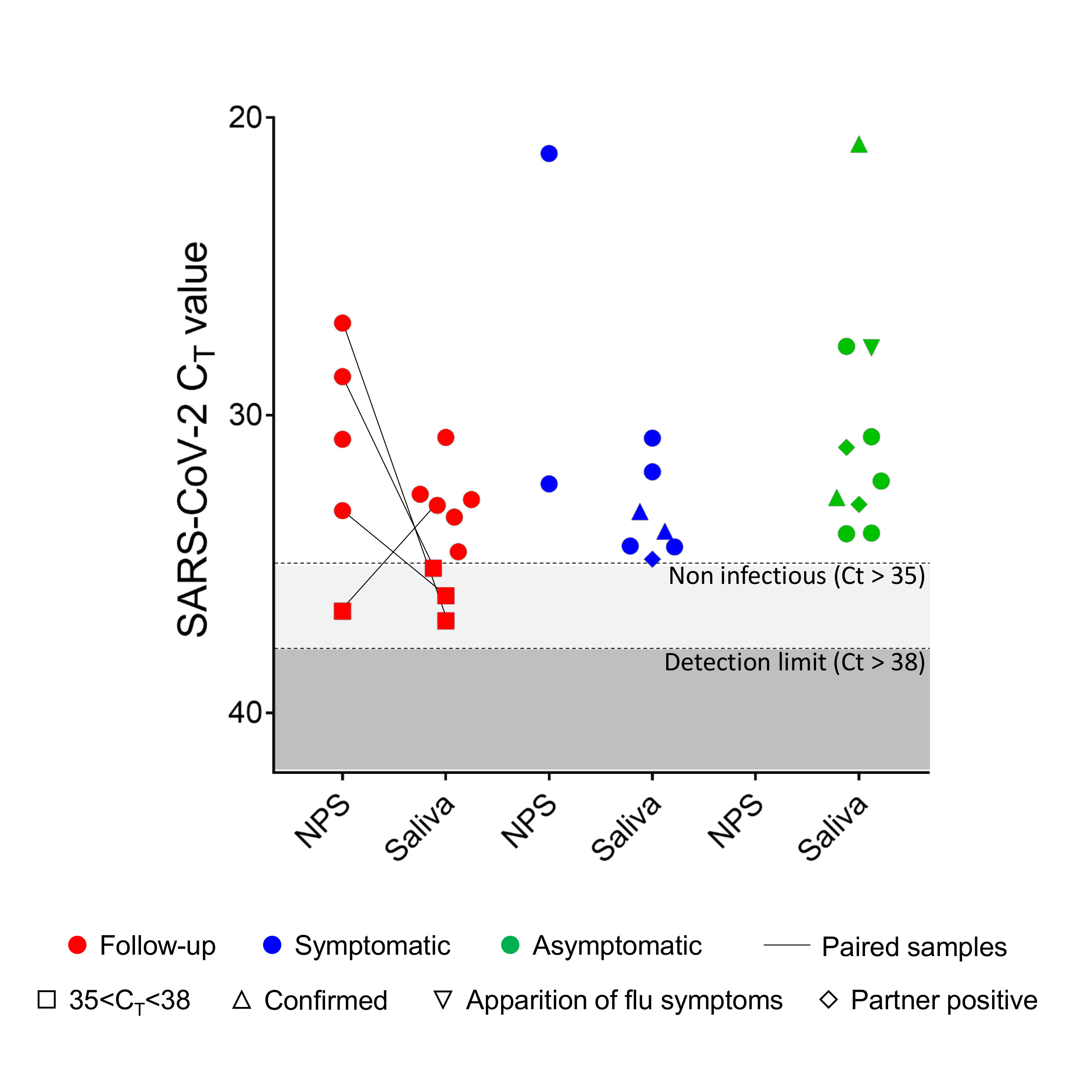

### Additional_File-12

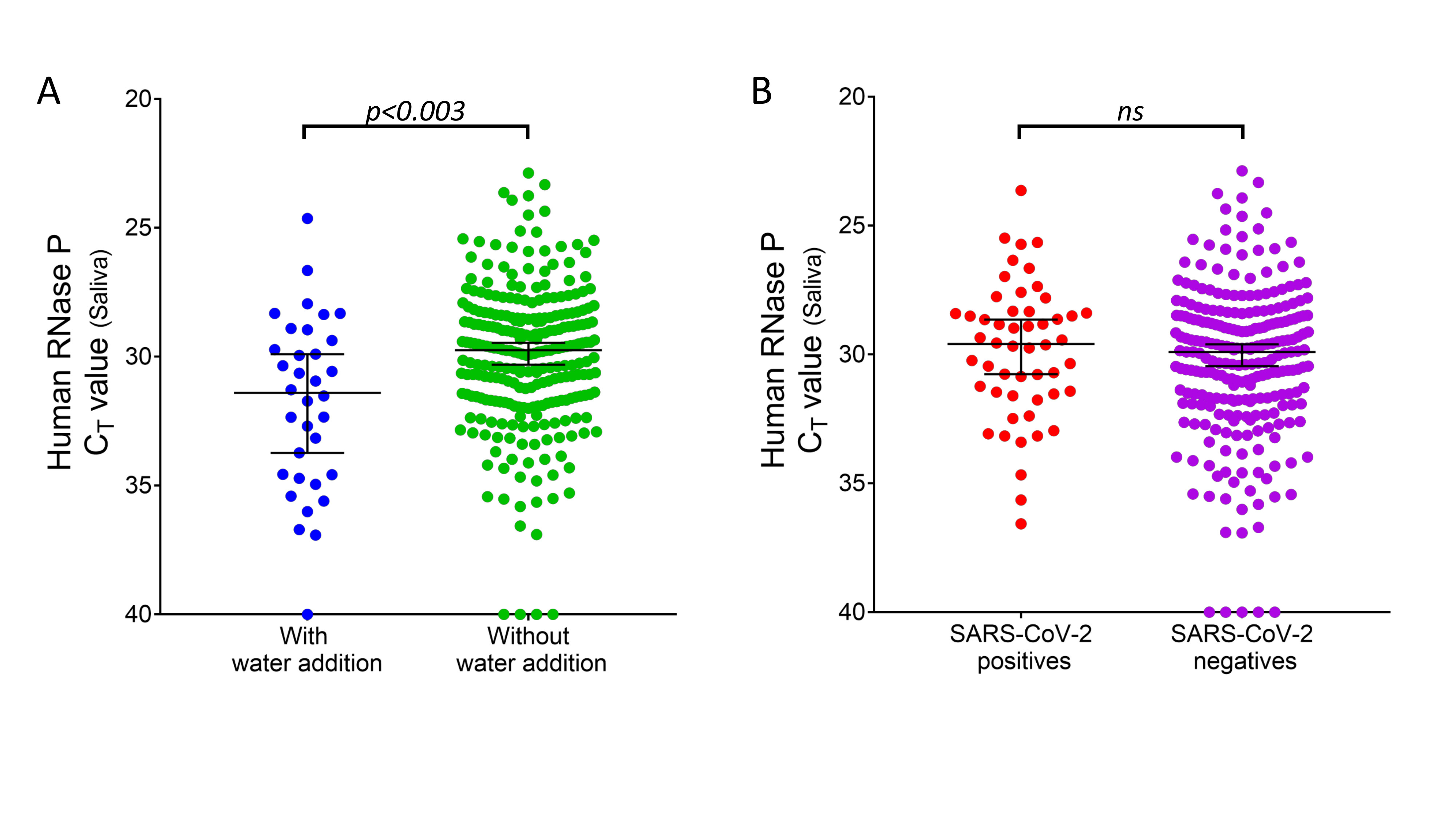
